# Long term follow-up of colorectal cancer screening attendees identifies differences in *Phascolarctobacterium spp*. using 16S rRNA and metagenome sequencing

**DOI:** 10.1101/2023.01.16.23284614

**Authors:** C. Bucher-Johannessen, E. Birkeland, E. Vinberg, V. Bemanian, G. Hoff, P. Berstad, TB. Rounge

## Abstract

**Background:** The microbiome has been implicated in the initiation and progression of colorectal cancer (CRC) in cross sectional studies. However, there is a lack of studies using prospectively collected samples.

**Methods:** We analysed 144 archived faecal samples from participants in the NORwegian Colorectal CAncer Prevention (NORCCAP) trial diagnosed with CRC or high-risk adenomas (HRA) at screening, or who remained cancer-free during 17 years of follow-up. We performed 16S rRNA sequencing of all samples, and metagenome sequencing on a subset of 47 samples. Differences in taxonomy and gene content between outcome groups were assessed for alpha and beta diversity, and differential abundance.

**Results:** Diversity and composition analyses showed no significant differences between CRC, HRA, and healthy controls. *Phascolarctobacterium succinatutens* was more abundant in CRC compared to healthy controls in both the 16S and metagenome data. The abundance of *Bifidobacterium* and *Lachnospiraceae spp*. were associated with time to CRC diagnosis.

**Conclusion:** Using a longitudinal study design, we identified three taxa as being potentially associated with CRC. These should be the focus of further studies of microbial changes occurring prior to CRC diagnosis.

## INTRODUCTION

CRC is the third most common cancer in men and the second in women world-wide [1, 2]. Symptoms are often unspecific, and many cases are detected at an advanced stage with reduced prospects for curative treatment. The progression towards CRC passes through stages of molecular and morphological changes from small and benign, through advanced adenoma, and finally to CRC. This adenoma-carcinoma sequence is estimated to take on average between 10-15 years [3]. This time window provides an opportunity to screen and potentially remove lesions that have not yet developed into clinical cancer and advanced stages [3, 4]. Several randomized studies have estimated that CRC screening by faecal tests reduces CRC mortality by 15-30% [5-8]. However, faecal based tests are hampered by both poor sensitivity and specificity, particularly for detection of CRC precursor lesions [9]. Therefore, there is a need for additional markers that can be used in faecal-based screening for CRC precursor lesions.

Analyses of the gut microbiome composition, diversity and functional potential have demonstrated that the gut microbiome of CRC patients is different from that of their healthy counterparts, making it a source of potential biomarkers for CRC [10-14]. The presence of certain microbes is strongly associated with CRC. The most frequently reported are *Fusobacterium nucleatum, Bacteroides fragilis* and pks+ *Escherichia coli*. Proposed mechanisms for a role of the microbiome in carcinogenesis include DNA damage through secretion of genotoxic compounds, induction of inflammation, and activation of pro-carcinogenic signalling pathways [15, 16]. While it has been shown that faecal tests in combination with microbial biomarkers are superior at separating healthy controls from CRC to that of a faecal test alone [17, 18], no specific bacterial profile is recognized as a biomarker for CRC. Still less is known about the role of the microbiome in the early stages of carcinogenesis.

To identify a pre-cancerous signal in the microbiome, there is a need for studies with sample collection prior to diagnosis and long-term follow-up. We performed microbiome sequencing on archived stool samples collected from screening attendees from the NORCCAP trial, with 17-year follow-up time after sigmoidoscopy screening. This study included both screening-detected cancers and CRC precursor lesions, as well as incident post-screening cancers, and healthy controls. We aimed at detecting community-wide and specific differences in the microbial profiles between CRC, HRA and healthy controls.

## METHOD

### Study design and participants

Details of the NORCCAP trial have been described previously [19-21]. Briefly, NORCCAP was a randomized clinical trial including 20,780 individuals offered sigmoidoscopy screening in the intervention arm and was performed in 1999-2000 (age group 55-64) and in 2001 (age group 50-54). The study recruited participants directly from the population registry of the Norwegian counties Oslo and Telemark. All participants were examined with flexible sigmoidoscopy, while 10,387 participants additionally delivered stool samples for an immunochemical faecal occult blood test (iFOBT – FlexSure OBT) and a fresh-frozen stool sample for biobanking. We selected a subset (n=300) of participants with archived fresh-frozen faecal samples for microbiome analyses (Fig. 1). Participants’ full CRC history was retrieved from the Cancer Registry of Norway in 2015 by using personal identification numbers and included the ICD-10 coded diagnoses C18, C19, and C20. Individuals with high-risk adenomas were defined as persons presenting with one or more adenomas of >=10 mm, with high grade dysplasia or villous components regardless of polyp size, or a person with three or more adenomas regardless of size, dysplasia and villosity. The control group was selected from a pool of participants with no findings (i.e., no lesions) at the screening examination (including low-risk adenomas) and who remained cancer-free during follow-up. Controls were selected by matching on sex, age, and examination date. This study received ethical approval from the Regional Committees for Medical and Health Research Ethics in South-Eastern Norway (ref: 22337).

**Figure 1.**
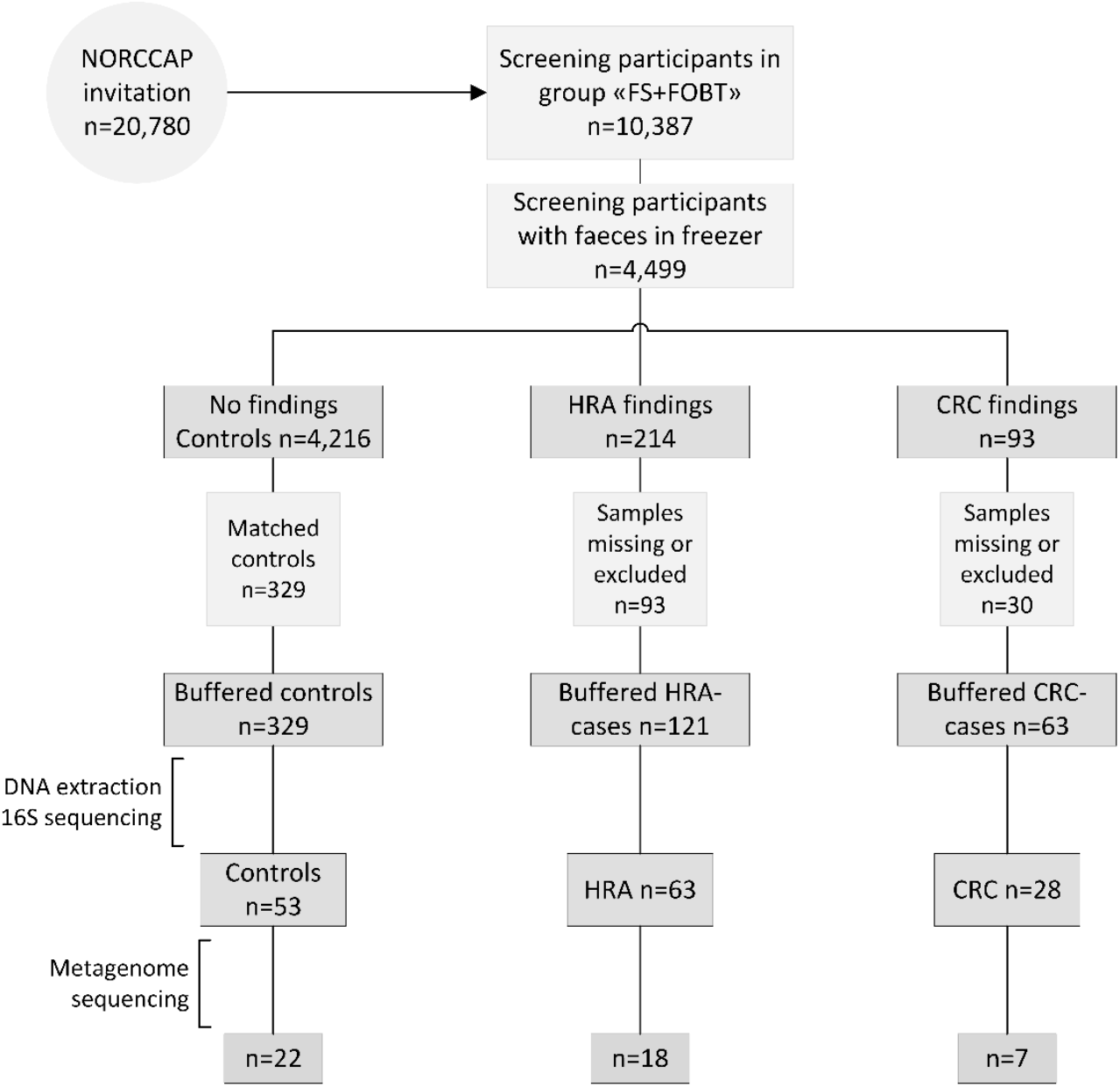
Recruitment flowchart. Half of the NORCCAP participants were invited to deliver a stool sample in addition to participating in sigmoidoscopy screening. Half of these faecal samples were stored below -20°C. A subset of samples diagnosed with CRC, HRA and matched controls were included in the study and homogenised in preservation buffers. Those with sufficient DNA extracted were included in 16S rRNA (n=144) and metagenome sequencing (n=47). FS fecal sigmoidoscopy, FOBT fecal occult blood test.

### DNA-extraction, library preparation and sequencing

Participants were asked to collect stool samples immediately after defecation at home in 20 mL vials and store the samples for at most seven days in freezer (−20 °C) before screening sigmoidoscopy. Samples were delivered to either of the two screening centres in Oslo or Telemark at time of sigmoidoscopy screening where further storage was at −20°C. We have previously demonstrated the feasibility of obtaining microbiota profiles from these archived stool samples [22]. Prior to DNA extraction, samples were thawed, homogenised and mixed with OMNIgene gut buffer. Extraction of DNA was carried out using the QIAsymphony automated extraction system, using the QIAsymphony DSP Virus/Pathogen Midikit (Qiagen, Hilden, Germany), after an off-board lysis protocol with some modifications. Each sample was lysed with bead-beating: a 500 μl sample aliquot was transferred to a Lysing Matrix E tube (MP Biomedicals) and mixed with 700 μl phosphate-buffered saline (PBS). The mixture was then shaken at 6.5 m/s for 45 seconds. After the bead-beating, 800 μl of the sample was mixed with 1055 μl of “off-board lysis buffer” (proteinase K, ATL buffer, ACL buffer and nuclease-free water) and incubated at 68°C for 15 min for lysis. Nucleic acid purification was performed on the QIAsymphony extraction robot using the Complex800_OBL_CR22796_ID 3489 protocol. Purified DNA was eluted in 60 μl AVE-buffer (Qiagen, Hilden, Germany). DNA purity was assessed using a Nanodrop2000 (Thermo Fisher Scientific, MA, USA), and the concentration was measured using a Qubit instrument (Thermo Fisher Scientific, MA, USA).

After DNA-extraction and sample quality assessment, libraries were prepared for 16S rRNA and shotgun metagenome sequencing. In total, 144 of available samples had sufficient DNA for 16S rRNA sequencing. Sample amplification was carried out using 16S primers S-D-Bact-0341-b-S-17 (5′CCTACGGGNGGCWGCAG′3) and S-D-Bact-0785-a-A-21 (5′GACTACHVGGGTATCTAATCC′ 3) to amplify the V3-V4 regions [23]. Amplification was performed using the Truseq (TS)-tailed1-step amplification protocol [24] with random spacers to shift the sequencing start. Paired-end 300 bp sequencing of PCR amplicons was performed on the Illumina Miseq instrument (Illumina, Inc. CA, USA) (Fig. S2A). Forty-seven of the samples had sufficient DNA for additional whole genome shotgun sequencing (Fig. S2B). The metagenomes provide additional taxonomical resolution and improved estimates of functional potential and were used for validation of the 16S rRNA sequencing results. Samples were cleaned up and concentrated using AMPure XP (Beckman Coulter, IN, USA) and normalized to a total input of 4 ng dsDNA. Sequencing libraries were prepared using the Riptide protocol (Twist Bioscience HQ, CA, USA), and sequenced on Illumina Novaseq paired end 2 × 130 bp. The Riptide protocol includes linear amplification with random primers and dideoxy nucleotide-induced self-termination, thereby avoiding DNA fragmentation [25]. Sequencing was performed at FIMM Technology Centre in Helsinki, Finland.

### Bioinformatics analyses

Initial quality control of 16S sequencing reads included removal of short reads (<50bp) and low-quality bases with average quality across four bases below 30 using Trimmomatic (v.0.35.2) [26]. Removal of primer sequences was done using Cutadapt (v.2020.2.0) [27] with the following options: forward primer: CCTACGGGNGGCWGCAG, reverse primer: GACTACHVGGGTATCTAATCC, primer error 0.1 and primer overlap 3. Fastqc and multiqc analyses were performed before and after trimming to ensure high quality of data [28]. Reads were imported into Qiime2 [29] and amplicon sequence variants (ASV) classification was performed using the Divisive Amplicon Denoising Algorithm 2 (DADA2) [30] plugin, including length trimming, merging, denoising and chimera removal. ASV classification was done using the SILVA 16S rRNA database (v.132) at a 97 percent similarity threshold [31]. ASV data were filtered for mitochondria and chloroplasts, and were rarefied to a depth of 9,000 reads for each sample. Metagenome functional profiles were predicted from the 16S data using Phylogenetic Investigation of Communities by Reconstruction of Unobserved States 2 (PICRUSt2) (v.2.3.0) with default settings, using rarefied count-tables as input, and mapping to MetaCyc database giving pathway abundance [32].

Metagenome reads were trimmed using Trimmomatic (v.0.66.0) [26] with a sliding window approach where reads with average quality across four bases below 30 or a read length of less than 30 basepairs were discarded. Following trimming, Bowtie2 (v.2.4.2) [33] and Samtools (v.1.12) [34] were used with default settings to remove reads mapping to the human genome. MetaPhlAn3 (v.3.0.4) was used for taxonomic classification with default parameters [35]. Percent abundances generated by MetaPhlAn3 were transformed into count-like tables by multiplying by the number of quality-trimmed reads per sample and dividing by 100. HUMAnN3 was used to profile genefamilies encoding microbial pathways (v.3.0.0.alpha.2), aggregating the data according to MetaCyc annotations using the UniRef90 (v.201901) database [35]. Pathway abundance data was corrected for sequencing depth by dividing by number of trimmed reads and multiplying by 10^6^.

### Statistical analysis

All statistical analyses were performed using R (v.3.5.3) and visualized using ggplot2 (v.3.3.2)[36]. To assess differences between CRC, HRA and the control group, statistical tests were made contrasting all three groups, or by combining CRC and HRAs. Additionally, analyses were performed within the CRC group, using time to diagnosis as the dependent variable. Differences between the three groups were evaluated using the Chi squared test for comparisons of two categorical variables, and Kruskal Wallis test (or, for two-group comparisons, Mann-Whitney U test) or Spearman’s correlation for comparisons of a continuous variable with a categorical and continuous variable, respectively. Statistical associations were considered significant at the p<0.05 level.

Microbial diversity was measured on ASV and species level for 16S and metagenome data, respectively. Alpha diversity was determined using richness, Shannon and Inverse Simpson indexes. Beta diversity was calculated using Bray-Curtis dissimilarity, all as implemented in the Phyloseq R package [37] (v.1.26.1). Associations between beta diversity and CRC, HRA and healthy controls were evaluated using permutational analysis of variance (PERMANOVA) with 999 permutations after adjustment for participant sex and screening centre, as implemented in the adonis function of the R package vegan (v.2.5-7).

Differential abundance analyses were performed independently on ASV/species, genus, phylum and pathways, and were adjusted for sex and screening centre. Before differential abundance analyses, we applied low abundance filtering, retaining all taxa/pathways with a read count of at least 10 in at least 10% of samples. Differential abundance analyses were performed using negative binominal model-based Wald test implemented in the DeSeq2 package with the type (poscounts) to account for the sparsity of microbiome data, and p-values were FDR adjusted to control for multiple testing (v.1.22.2) [38].

## RESULTS

### Study population

Stool samples from 144 NORCCAP screening participants were selected for 16S sequencing based on registry follow-up data and initial screening results. Metagenome sequencing was also performed on 47 of these with the highest DNA amounts. All 144 participants in this study underwent sigmoidoscopy. Five cases of CRC (3.5%) were detected during screening. Based on registry follow-up, 23 (16%) participants received a CRC diagnosis within 17 years after screening (Fig. 2, Table 1). The median time from screening to CRC diagnosis was 7.4 years (range 0-16 years) and the median age at CRC diagnosis was 65.7 years (range 54-77), including both screening-detected and follow-up diagnosed CRC. Other screening-detected lesions included 63 HRAs (44% of study participants). Fifty-three (37%) participants had no findings of adenomas or CRC during sigmoidoscopy and were cancer free during follow-up; these constituted the control group. The median age for all groups at sample collection was 57 years (range 51-65). We observed a significantly different distribution of sex and screening centre between CRC, HRA and healthy controls (p<0.05). In total, 87 (60%) samples were from male participants and 89 (62%) samples were from the Telemark screening centre.

**Figure 2.**
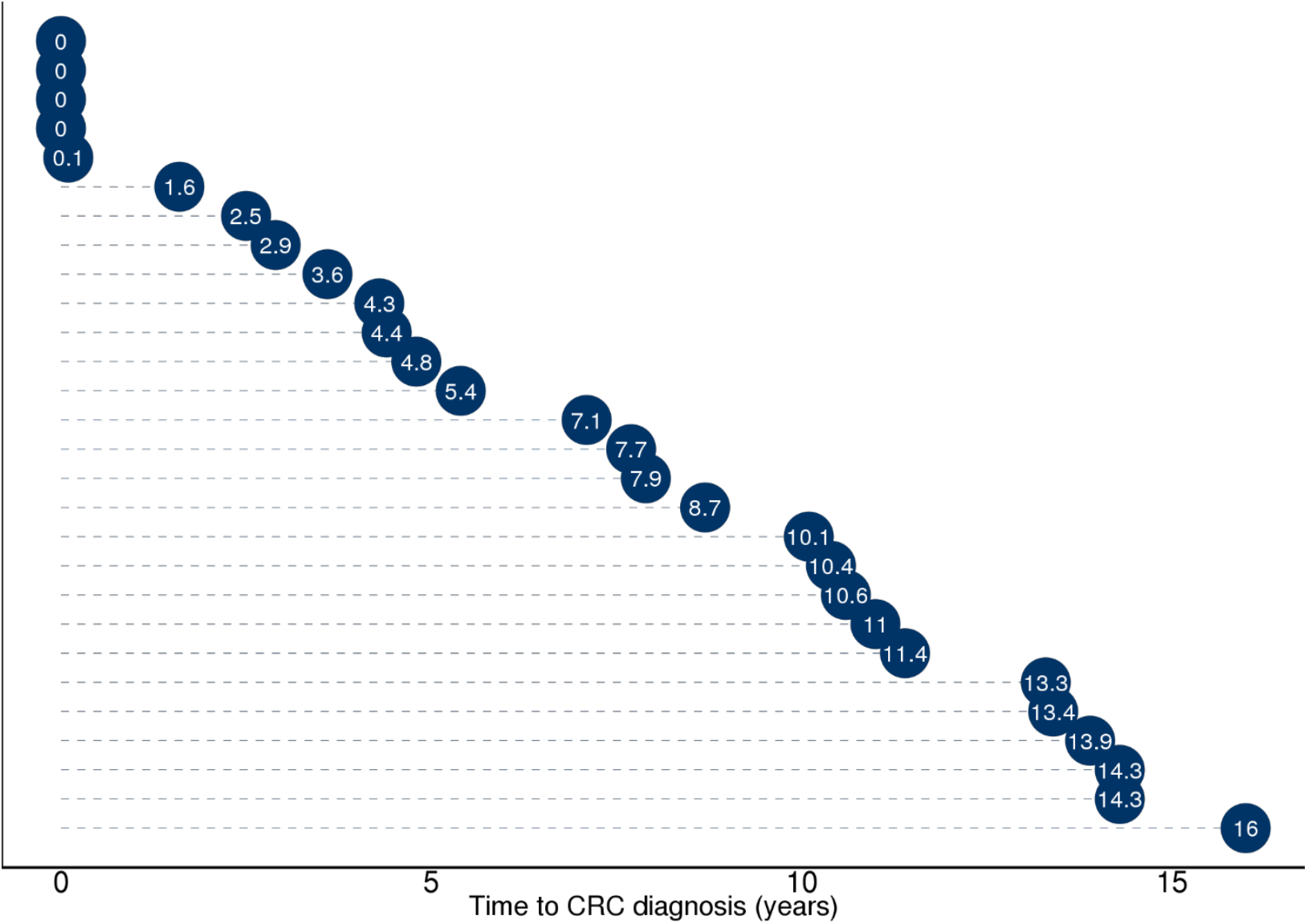
Overview of time from sample collection to cancer diagnosis for the 23 study participants who received a CRC diagnosis during screening or follow-up. Five participants with time to diagnosis ≤ 0.1 years received the diagnosis during screening.

**Table 1.**
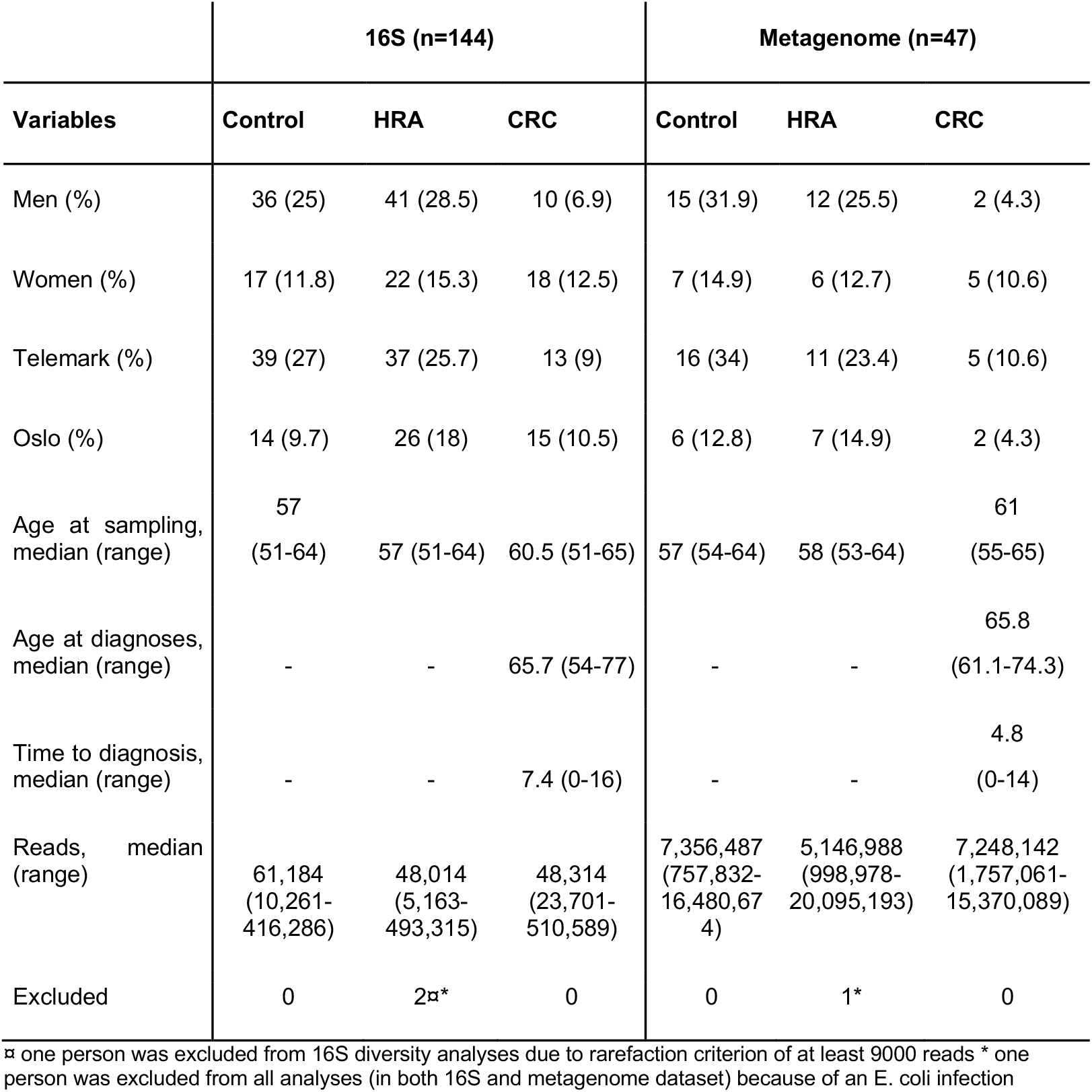
Characteristics of study participants and samples

### Gut microbiome diversity

16S sequencing of 144 samples generated 11.8 million trimmed reads with a median read depth per sample of 50,205 (range 5,163-510,589). We identified in total 7,228 ASVs mapped to 337 species, 229 genus and 18 phyla. The median number of observed ASVs was 213.5 (range 79-603). Metagenomic sequencing of 47 samples resulted in 361million trimmed reads with a median read depth of 6.2 million reads (0.76-20.1). In total, 561 taxa were identified, including 323 species, 116 genus and 8 phyla. The median number of species per sample was 73 (34-107). ASV distribution for individual samples showed one sample with 83% of reads belonging to two ASVs within the genus *Escherichia-Shigella*. This was confirmed in the metagenome data where 95% of reads belonged to the species *Escherichia coli*. As this indicated an unrelated acute infection, the sample was excluded from further analyses (Fig. S1).

Rarefying 16S data to 9,000 reads resulted in exclusion of one sample with lower sequencing coverage, leaving 142 samples for 16S diversity analyses. Forty-six samples were used for metagenome diversity analyses. We found no significant differences in alpha (unadjusted) or beta diversity of taxa or pathways between CRC, HRA and healthy controls (Fig. 3A-D, 4A-D, p>0.05 for all comparisons). This finding remained consistent when grouping CRC and HRA cases together, when looking at time to diagnosis, when considering metagenome data, and when adjusting for sex and screening centre.

**Figure 3.**
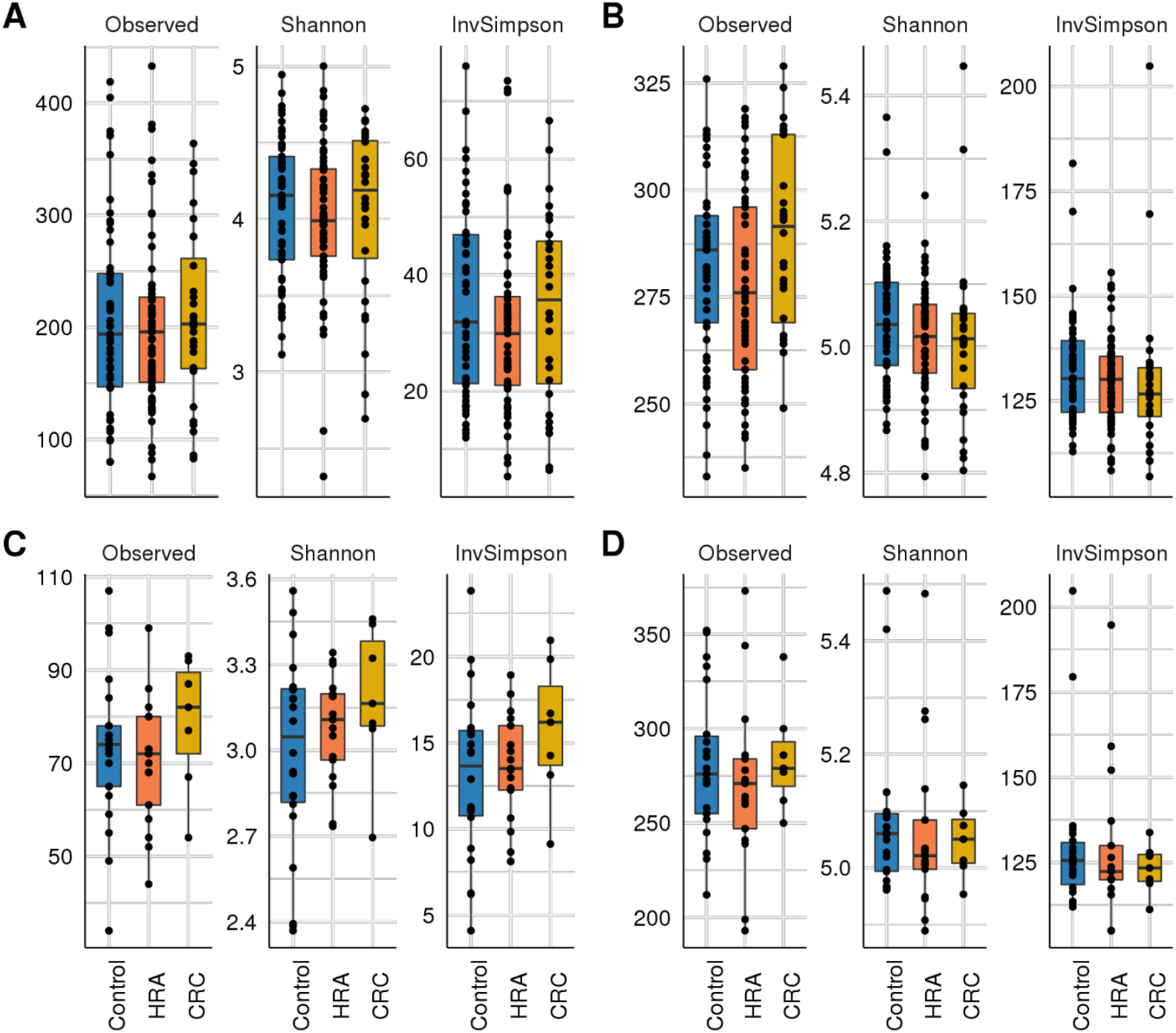
Alpha diversity: box plots with taxa/pathway richness (Observed), and Shannon and Inverse Simpson (InvSimpson) diversity indices in CRC, HRA and controls for A) amplicon sequence variants from 16S sequencing data B) estimated MetaCyc pathways derived from 16S data C) Species abundance based on metagenome shotgun sequencing D) Metacyc pathways based on metagenome shotgun sequencing.

**Figure 4.**
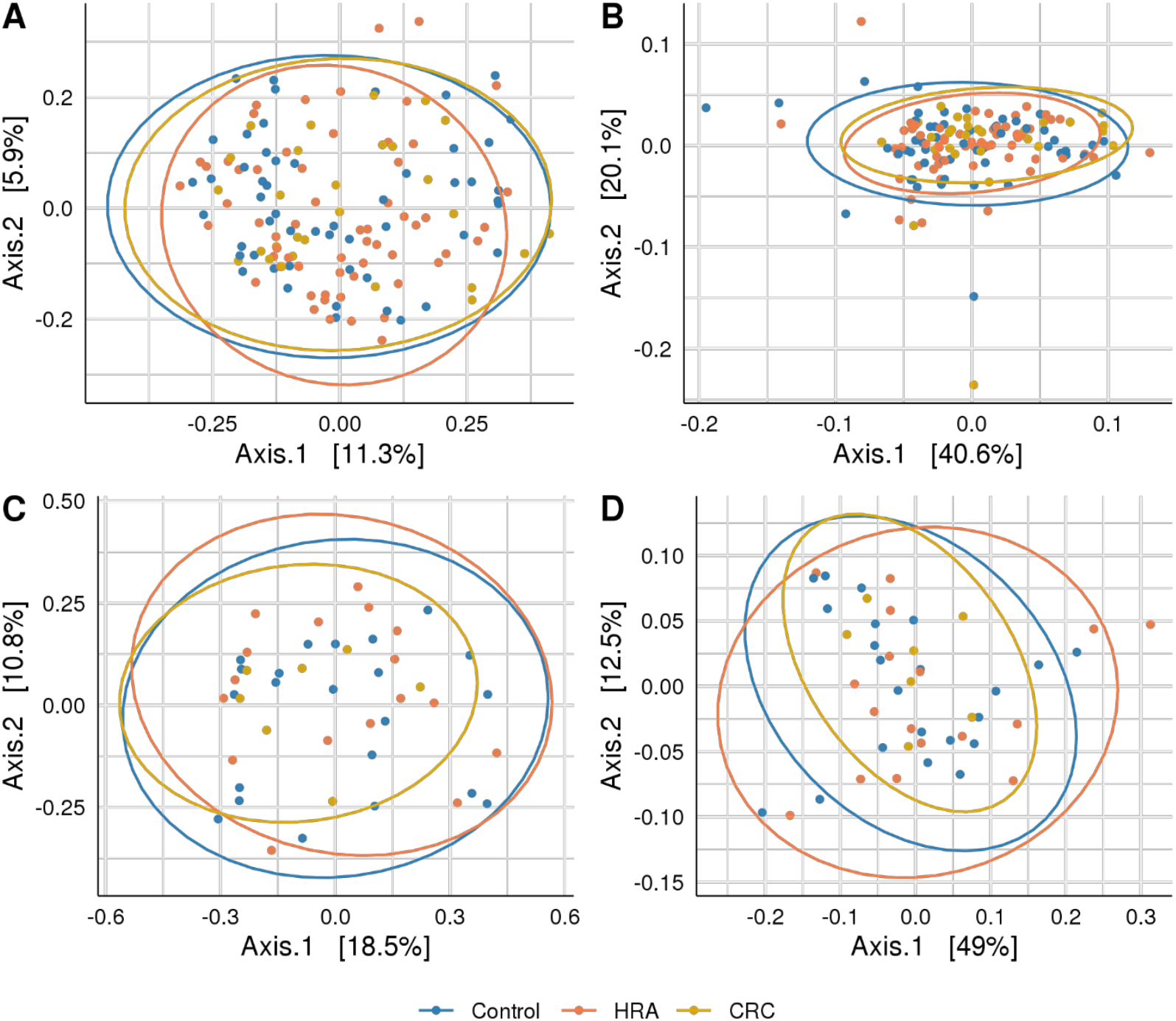
Beta diversity: PCoA plots with Bray Curtis dissimilarity indices between CRC, HRA and controls for A) amplicon sequence variants from 16S sequencing data B) MetaCyc pathways derived from 16S sequencing data C) Species abundance based on metagenome shotgun sequencing D) Metacyc pathways based on metagenome shotgun sequencing. Ellipses describe 95% of group variation for the principal coordinate axes.

### Differentially abundant taxa and pathways

We evaluated differences in abundance of individual taxa and pathways between the outcome groups using the abundance of ASV/species, genus, phylum or pathways. We further assessed associations of ASVs with the time elapsed from sample collection to CRC diagnosis.

#### CRC vs. Control

For the 16S data, the ASV *Phascolarctobacterium uncultured bacterium* and the phylum *Firmicutes* were significantly more abundant in CRC than controls (FDR p<0.05, Table 2 and Fig. 5A). Similarly, in the metagenome data *Phascolarctobacterium succinatutens* was significantly more abundant in CRC. For the metagenome data, in total 9 species were differentially abundant (FDR p<0.05, Table 2 and Fig. 5C). Five of these were significantly higher in CRC compared to control, whereas 4 were significantly lower. The genus *Acidaminococcus* was significantly higher in CRC. Four pathways were significantly lower in CRC compared to controls.

**Table 2.**
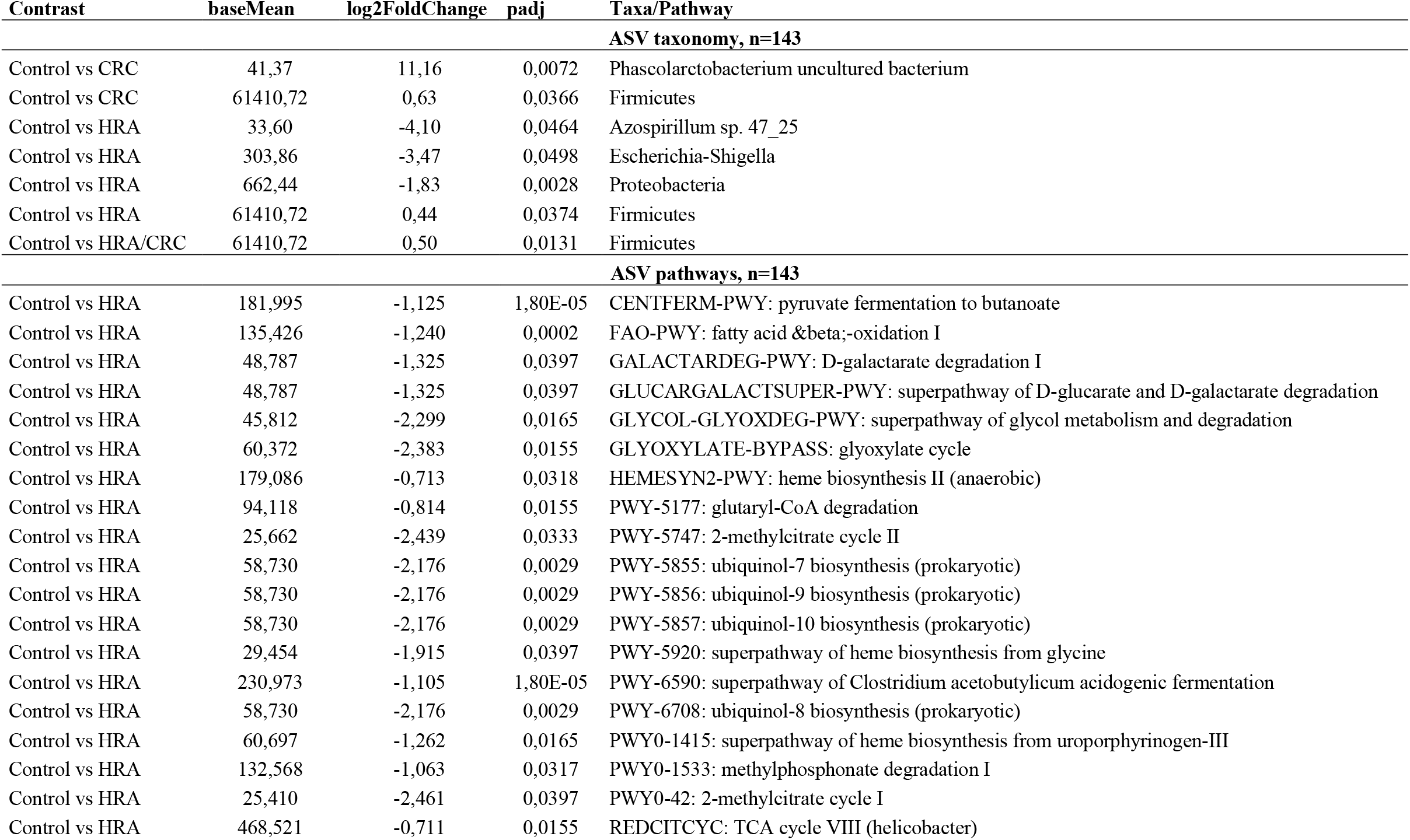
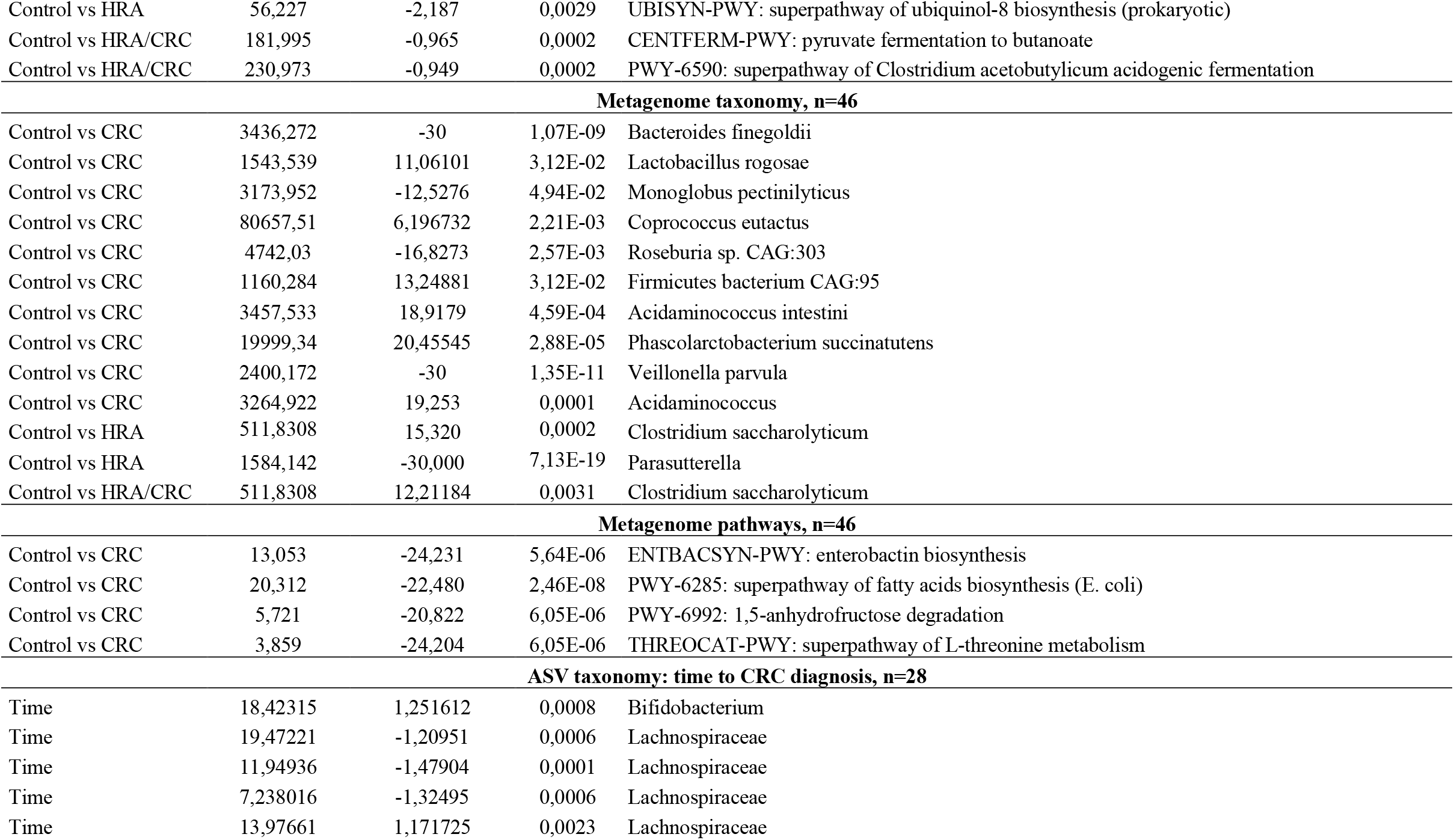
Differential abundance aalyses of taxa and pathways between CRC, HRA and healthy controls. Log-2foldchange indicates the magnitude and direction of difference in abundance. Analyses were adjusted for sex and screening centre.

| Contrast | baseMean | log2FoldChange | padj | Taxa/Pathway |
| --- | --- | --- | --- | --- |
| **ASV taxonomy, n=143** |  |  |  |  |
| Control vs CRC | 41,37 | 11,16 | 0,0072 | Phascolarctobacterium uncultured bacterium |
| Control vs CRC | 61410,72 | 0,63 | 0,0366 | Firmicutes |
| Control vs HRA | 33,60 | -4,10 | 0,0464 | Azospirillum sp. 47_25 |
| Control vs HRA | 303,86 | -3,47 | 0,0498 | Escherichia-Shigella |
| Control vs HRA | 662,44 | -1,83 | 0,0028 | Proteobacteria |
| Control vs HRA | 61410,72 | 0,44 | 0,0374 | Firmicutes |
| Control vs HRA/CRC | 61410,72 | 0,50 | 0,0131 | Firmicutes |
| **ASV pathways, n=143** |  |  |  |  |
| Control vs HRA | 181,995 | -1,125 | 1,80E-05 | CENTFERM-PWY: pyruvate fermentation to butanoate |
| Control vs HRA | 135,426 | -1,240 | 0,0002 | FAO-PWY: fatty acid & beta;-oxidation I |
| Control vs HRA | 48,787 | -1,325 | 0,0397 | GALACTARDEG-PWY: D-galactarate degradation I |
| Control vs HRA | 48,787 | -1,325 | 0,0397 | GLUCARGALACTSUPER-PWY: superpathway of D-glucarate and D-galactarate degradation |
| Control vs HRA | 45,812 | -2,299 | 0,0165 | GLYCOL-GLYOXDEG-PWY: superpathway of glycol metabolism and degradation |
| Control vs HRA | 60,372 | -2,383 | 0,0155 | GLYOXYLATE-BYPASS: glyoxylate cycle |
| Control vs HRA | 179,086 | -0,713 | 0,0318 | HEMESYN2-PWY: heme biosynthesis II (anaerobic) |
| Control vs HRA | 94,118 | -0,814 | 0,0155 | PWY-5177: glutaryl-CoA degradation |
| Control vs HRA | 25,662 | -2,439 | 0,0333 | PWY-5747: 2-methylcitrate cycle II |
| Control vs HRA | 58,730 | -2,176 | 0,0029 | PWY-5855: ubiquinol-7 biosynthesis (prokaryotic) |
| Control vs HRA | 58,730 | -2,176 | 0,0029 | PWY-5856: ubiquinol-9 biosynthesis (prokaryotic) |
| Control vs HRA | 58,730 | -2,176 | 0,0029 | PWY-5857: ubiquinol-10 biosynthesis (prokaryotic) |
| Control vs HRA | 29,454 | -1,915 | 0,0397 | PWY-5920: superpathway of heme biosynthesis from glycine |
| Control vs HRA | 230,973 | -1,105 | 1,80E-05 | PWY-6590: superpathway of Clostridium acetobutylicum acidogenic fermentation |
| Control vs HRA | 58,730 | -2,176 | 0,0029 | PWY-6708: ubiquinol-8 biosynthesis (prokaryotic) |
| Control vs HRA | 60,697 | -1,262 | 0,0165 | PWY0-1415: superpathway of heme biosynthesis from uroporphyrinogen-III |
| Control vs HRA | 132,568 | -1,063 | 0,0317 | PWY0-1533: methylphosphonate degradation I |
| Control vs HRA | 25,410 | -2,461 | 0,0397 | PWY0-42: 2-methylcitrate cycle I |
| Control vs HRA | 468,521 | -0,711 | 0,0155 | REDCITCYC: TCA cycle VIII (helicobacter) |
| Control vs HRA | 56,227 | -2,187 | 0,0029 | UBISYN-PWY: superpathway of ubiquinol-8 biosynthesis (prokaryotic) |
| Control vs HRA/CRC | 181,995 | -0,965 | 0,0002 | CENTFERM-PWY: pyruvate fermentation to butanoate |
| Control vs HRA/CRC | 230,973 | -0,949 | 0,0002 | PWY-6590: superpathway of Clostridium acetobutylicum acidogenic fermentation |
| Metagenome taxonomy, n=46 |  |  |  |  |
| Control vs CRC | 3436,272 | -30 | 1,07E-09 | Bacteroides finegoldii |
| Control vs CRC | 1543,539 | 11,06101 | 3,12E-02 | Lactobacillus rogosae |
| Control vs CRC | 3173,952 | -12,5276 | 4,94E-02 | Monoglobus pectinilyticus |
| Control vs CRC | 80657,51 | 6,196732 | 2,21E-03 | Coprococcus eutactus |
| Control vs CRC | 4742,03 | -16,8273 | 2,57E-03 | Roseburia sp. CAG:303 |
| Control vs CRC | 1160,284 | 13,24881 | 3,12E-02 | Firmicutes bacterium CAG:95 |
| Control vs CRC | 3457,533 | 18,9179 | 4,59E-04 | Acidaminococcus intestini |
| Control vs CRC | 19999,34 | 20,45545 | 2,88E-05 | Phascolarctobacterium succinatutens |
| Control vs CRC | 2400,172 | -30 | 1,35E-11 | Veillonella parvula |
| Control vs CRC | 3264,922 | 19,253 | 0,0001 | Acidaminococcus |
| Control vs HRA | 511,8308 | 15,320 | 0,0002 | Clostridium saccharolyticum |
| Control vs HRA | 1584,142 | -30,000 | 7,13E-19 | Parasutterella |
| Control vs HRA/CRC | 511,8308 | 12,21184 | 0,0031 | Clostridium saccharolyticum |
| Metagenome pathways, n=46 |  |  |  |  |
| Control vs CRC | 13,053 | -24,231 | 5,64E-06 | ENTBACSYN-PWY: enterobactin biosynthesis |
| Control vs CRC | 20,312 | -22,480 | 2,46E-08 | PWY-6285: superpathway of fatty acids biosynthesis (E. coli) |
| Control vs CRC | 5,721 | -20,822 | 6,05E-06 | PWY-6992: 1,5-anhydrofructose degradation |
| Control vs CRC | 3,859 | -24,204 | 6,05E-06 | THREOCAT-PWY: superpathway of L-threonine metabolism |
| ASV taxonomy: time to CRC diagnosis, n=28 |  |  |  |  |
| Time | 18,42315 | 1,251612 | 0,0008 | Bifidobacterium |
| Time | 19,47221 | -1,20951 | 0,0006 | Lachnospiraceae |
| Time | 11,94936 | -1,47904 | 0,0001 | Lachnospiraceae |
| Time | 7,238016 | -1,32495 | 0,0006 | Lachnospiraceae |
| Time | 13,97661 | 1,171725 | 0,0023 | Lachnospiraceae |

**Figure 5.**
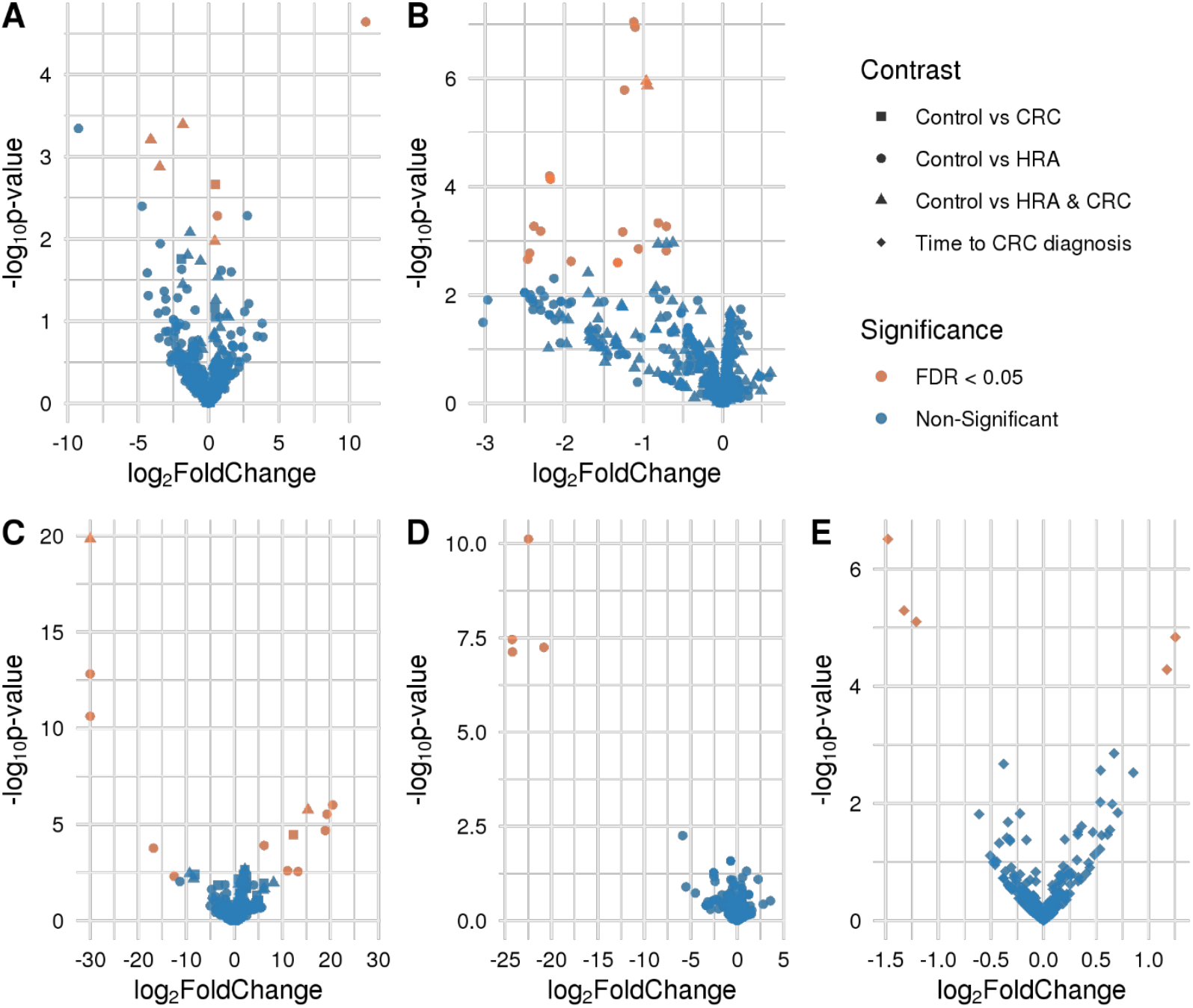
Volcano plots showing differences in abundance of taxa and pathways between groups. FDR-significant differentially abundant taxa or pathways are highlighted in red. Group comparisons are indicated by different shapes where the control group or a shorter time to diagnosis is considered the reference. Differential abundance was analysed for A) amplicon sequence variants from 16S sequencing data B) MetaCyc pathways derived from 16S sequencing data C) Species abundance based on metagenome shotgun sequencing D) Metacyc pathways based on metagenome shotgun sequencing E) amplicon sequence variants from 16S sequencing data and time to CRC diagnosis.

#### HRA vs. Control

For 16S data, the genera *Azospirillum sp. 47_25* and *Escherichia-Shigella* were lower in HRA compared to controls (FDR p<0.05, Table 2 and Fig. 5A). The phyla *Proteobacteria* and *Firmicutes* were lower and higher in HRA compared to controls, respectively. The direction of differences for these phyla was similar in the metagenome data, though not significant. Twenty pathways were lower in HRA based on 16S data. Of these, three pathways were related to heme biosynthesis: HEMESYN2-PWY (heme biosynthesis II (anaerobic)), PWY-5920 (superpathway of heme biosynthesis from glycine), and PWY0-1415 (superpathway of heme biosynthesis from uroporphyrinogen-III) (FDR p<0.05, Table 2 and Fig. 5B). The direction was similar for PWY0-1415 in the metagenome data. We also observed differences in REDCITCYC (TCA cycle VIII (helicobacter)) and the closely related pathways PWY0-42 (methylcitrate cycle I), PWY-5747 (methylcitrate cycle II) and GLYOXYLATE-BYPASS (glyoxylate cycle). For metagenome data the species *Clostridium saccharolyticum* was significantly higher and genus *Parasutterella* was significantly lower in HRA compared to controls (FDR p<0.05, Table 2 and Fig. 5C).

#### HRA&CRC vs. Control

For 16S, when considering HRA and CRC as one group and comparing it to controls, the phylum *Firmicutes* was significantly higher in HRA/CRC (FDR p<0.05, Table 2 and Fig. 5A). The same non-significant trend was observed in the metagenome data. Pathways CENTFERM-PWY (pyruvate fermentation to butanoate) and PWY-6590 (superpathway of Clostridium acetobutylicum acidogenic fermentation) were lower in HRA/CRC (FDR p<0.05, Table 2 and Fig. 5B). For metagenome data, the species *Clostridium saccharolyticum* was significantly more abundant in the HRA/CRC group (FDR p<0.05, Table 2 and Fig. 5C).

#### Time to diagnosis

Assessing the CRC group only, those with a longer interval between sample collection and diagnosis had higher abundance of one genus, *Bifidobacterium* and one ASV within the *Lachnospiraceae* family. Additionally, three ASVs within *Lachnospiraceae* were lower in those with a long time to diagnosis (FDR p<0.05, Fig. 6 and Table 2).

## DISCUSSION

Using both 16S rRNA and metagenome sequencing data, we analysed the microbial differences between CRC, HRA and healthy controls of 144 screening attendees with long-term follow-up data. *Phascolarctobacterium spp*., were more abundant in the CRC compared to controls and four ASVs belonging to the *Lachnospiraceae* family, and *Bifidobacterium* were associated with time to CRC diagnosis. Several heme biosynthesis pathways were less abundant in HRA. We did not observe compositional differences between CRC, HRA and healthy controls, and identified no correlation between richness and time to diagnosis in the CRC group.

We identified *Phascolarctobacterium uncultured bacterium* and *Phascolarctobacterium succinatutens* in the 16S and metagenome data respectively, as being significantly higher in CRC compared to healthy controls. These annotations likely represent the same species. Three studies have reported similar findings [39-41]. Interestingly, [41] et al. found an elevation in *P. succinatutens* in the early stages of CRC, from polypoid adenomas to stage 1 CRC. *P. succinatutens* is broadly distributed in the GI tract and converts succinate into propionate. The strain can likely not ferment any other short-chain fatty acids or carbohydrates [42]. Succinate is a tricarboxylic acid (TCA) cycle intermediate and is produced both by the host and the microbiota, including CRC-associated bacteria *B. fragilis* and *F. nucleatum*. Increased succinate in the colon has been linked to gut inflammation and disease, while increased propionate is thought to be anti-inflammatory [43, 44]. Succinate is proposed to mediate cross-talk as a signaling metabolite that act as a positive regulator of intestinal gluconeogenesis [44, 45] and thermogenesis [46] We also report several pathways related to the TCA-cycle to be lower in HRA compared to controls. [47] et al found this pathway to be increased in cancer.

Three pathways related to heme biosynthesis were significantly lower in the HRA group compared to controls. While heme uptake, biosynthesis and export in bacteria are not fully understood [48, 49], bleeding tumours release heme into the gut lumen. This might create a niche for heme scavenging bacteria which could outcompete those who rely on heme biosynthesis.

*Bifidobacterium* and four ASVs belonging to *Lachnospiraceaecea* family were associated with time to diagnosis. Bifidobacterium is a lactic acid producing bacteria, aiding in colonocyte renewal and inhibiting growth of pathogens. Two studies found Bifidobacterium to be lower in persons with lesions compared to controls [3, 50]. This is in line with our findings that lower levels are associated with a shorter time to diagnosis. We observed different members of the *Lachnospiraceaecea* family showing diverging associations with time to diagnosis. This family was found to be enriched in controls compared to patients with lesions [51]. Some members of the *Lachnospiraceaecea* family can produce the short-chain fatty acid butyrate [52]. Butyrate aids in cell renewal of colonocytes, serves as a carbon source for the TCA cycle, and has anti-inflammatory and anti-tumorigenic properties [53, 54].

We found no difference in diversity or composition between CRC, HRA and controls. Results from similar studies seem to be conflicting, both for diversity and composition analyses. [11, 55-58]. Smaller differences in the microbiome of adenomas and healthy controls have been observed than those observed between cancers and healthy controls [3, 11]. Unlike previous studies in the field, many of our samples were collected from asymptomatic subjects, years before diagnosis of cancer. While our results indicate no overall difference in diversity or composition, it is possible that we have been underpowered or that factors related to study design and technical challenges have led us to miss any small differences in these ecological measures.

This study has some other noteworthy limitations. Firstly, our samples were stored for 17 years and could possibly be degraded. We do know that these samples have few freeze-thaw cycles [22], but they were stored without a stabilizing agent which could to some extent influence the composition of faecal samples [59, 60]. Further, we lack information on important confounding factors such as diet, lifestyle factors, body mass index, and antibiotics use affecting microbiome composition [53, 61]. Lastly, we observed a high abundance of the phylum *Firmicutes* in our 16S data, but a similar composition was not observed for the metagenome data. This is likely due to the choice of primers, where for marker gene studies, certain primers favour the amplification of specific taxa [62]. Still, this did likely not affect the differential abundance analyses, as the bias was uniform across samples.

## Conclusions

The present study is, to the best of our knowledge, the first to examine gut microbiome samples collected several years prior to CRC diagnosis. We did not find any differences between diversity and composition of the gut microbiome and the presence of CRC, HRA, and controls. However, analyses identified several taxa and pathways that were differentially abundant. Our study found that the succinate-metabolising, associated with inflammation, *Phascolarctobacterium succinatutens* was more prevalent in individuals diagnosed with CRC than in healthy controls, identified using both 16S and metagenome data. In this population-based screening setting we also show that CRC-associated taxa are identifiable years prior to diagnosis of CRC.

## Data Availability

The datasets generated and/or analysed during the current study are not publicly available because this is human data and individuals privacy could be compromised but data is available from the corresponding author on reasonable request.

## Declarations

### Ethics approval and consent to participate

This study received ethical approval from the Regional Committees for Medical and Health Research Ethics in South-Eastern Norway (ref: 22337).

### Consent for publication

Not applicable

### Availability of data and materials

The datasets generated and/or analysed during the current study are not publicly available because this is human data and individuals’ privacy could be compromised but data is available from the corresponding author on reasonable request.

### Authors’ contributions

TBR and GH designed the study. CBJ and EB analysed the data. EV and VB performed sample preparation and lab work. CBJ, EB and TBR drafted the manuscript. All authors commented and approved the final manuscript.

## Acknowledgements

We would like to thank Jan Inge Nordby for his contribution on sample preparation and lab work. Library preparation and sequencing was performed at FIMM Technology Centre supported by HiLIFE and Biocenter Finland. Especially, we would like to thank Tiina Hannunen, Harri A. Kangas and Pekka J. Ellonen for the good cooperation, service and communication. We would also like to acknowledge Even Sannes Riiser for his early contributions on lab coordination and bioinformatics analyses. Finally, this article is a result of cooperations and scientific discussions among colleagues in our research group. Therefore, we want to thank Ane Sørlie Kværner, Ekaterina Avershina, Paula Istvan and Maja Jacobsen.

