## Supplementary information for "Long term follow-up of colorectal cancer screening attendees identifies differences in *Phascolarctobacterium spp*. using 16S rRNA and metagenome sequencing"

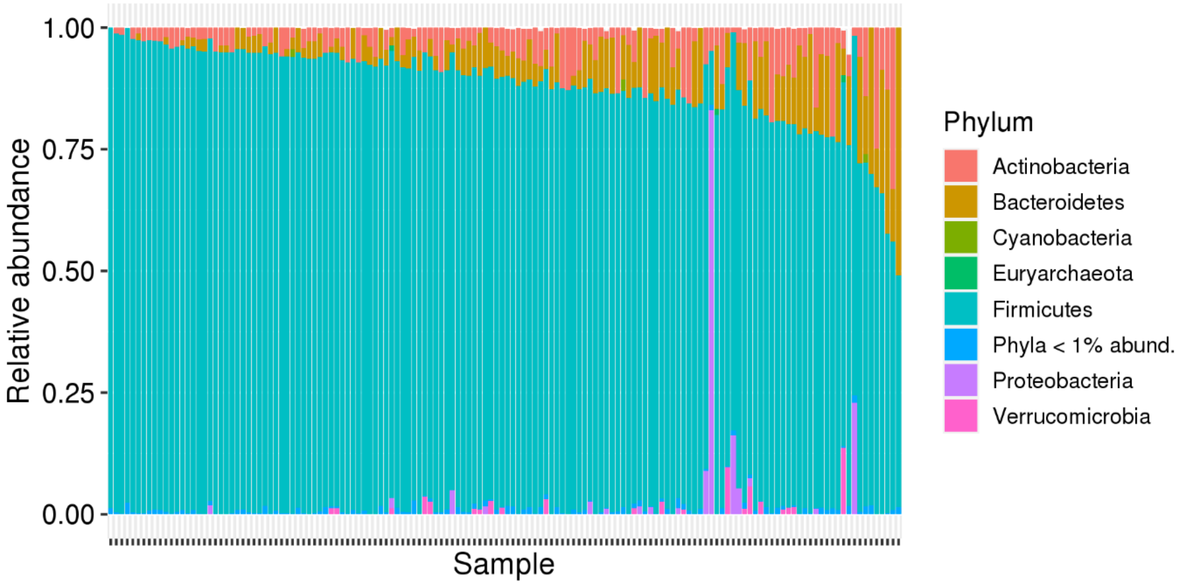


Figure S1 Relative abundance plot on phylum level for all samples from the 16S data.

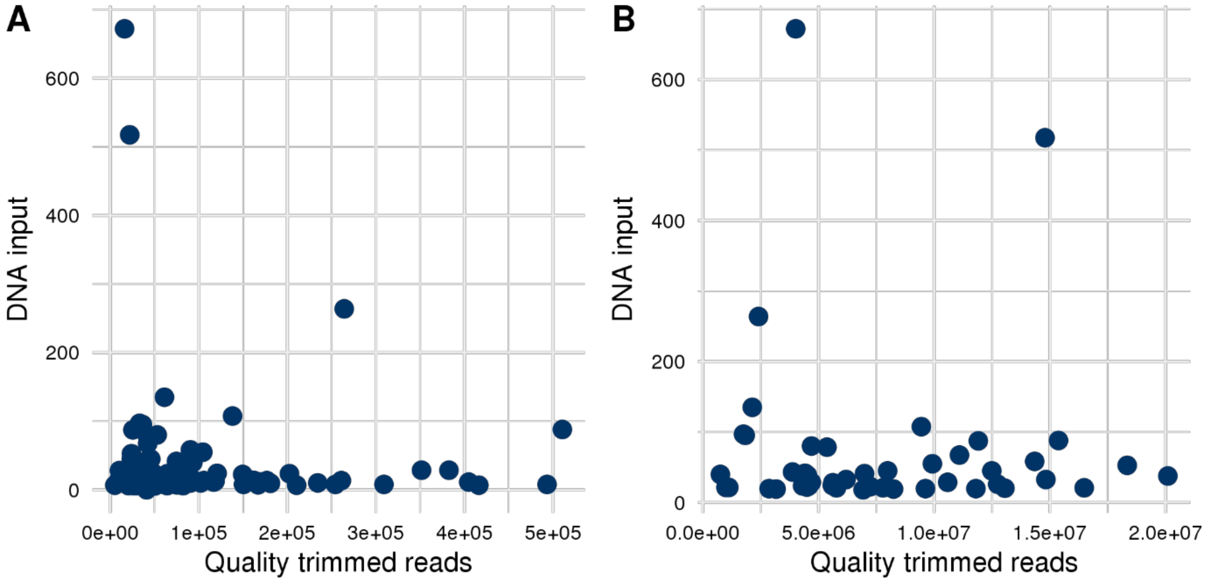


Figure 2 Quality trimmed reads plotted against DNA concentration after DNA isolation for the A) 16S data B) Metagenome data
